# Accelerating Medicines Partnership: Parkinson’s Disease. Genetic Resource

**DOI:** 10.1101/2020.11.19.20235192

**Authors:** Hirotaka Iwaki, Hampton L. Leonard, Mary B. Makarious, Matt Bookman, Barry Landin, David Vismer, Bradford Casey, J. Raphael Gibbs, Dena G. Hernandez, Cornelis Blauwendraat, Daniel Vitale, Yeajin Song, Dinesh Kumar, Clifton L. Dalgard, Mahdiar Sadeghi, Xianjun Dong, Leonie Misquitta, Sonja W. Scholz, Clemens R. Scherzer, Mike A. Nalls, Shameek Biswas, Andrew B Singleton, Uniformed Services University of the Health Sciences Associates, AMP PD Whole Genome Sequencing Working Group, AMP PD consortium

**Affiliations:** Data Tecnica International, Glen Echo, MD, USA; Center for Alzheimer’s and Related Dementias, National Institute on Aging, Bethesda, MD, USA; Laboratory of Neurogenetics, National Institute on Aging, Bethesda, MD, USA; Verily Life Sciences, San Jose, CA, USA; Technome, Herndon, VA, USA; The Michael J. Fox Foundation for Parkinson’s Research, New York, NY, USA; Sanofi, Seattle, WA, USA; Department of Anatomy, Physiology & Genetics, Uniformed Services University of the Health Sciences, Bethesda, MD, USA; The American Genome Center, Uniformed Services University of the Health Sciences, Bethesda, MD, USA; Northeastern University, Boston, MA, USA; Harvard Medical School, Brigham and Women’s Hospital, Boston, MA, USA; Publicis Sapient, Bethesda, MD, USA; National Institute of Neurological Disorders and Stroke, Bethesda, MD, USA; Department of Neurology, Johns Hopkins University, Baltimore, MD, USA; Bristol Myers Squibb, Seattle, WA, USA

## Abstract

**Background:** Whole-genome sequencing (WGS) data is available from several large studies across a variety of diseases and traits. However, massive storage and computation resources are required to use these data, and, to achieve the sufficient power for discoveries, harmonization of multiple cohorts is critical.

**Objectives:** The Accelerating Medicines Partnership Parkinson’s Disease (AMP PD) program has developed a research platform for Parkinson’s disease (PD) which integrates the storage and analysis of WGS data, RNA expression data, and clinical data, harmonized across multiple cohort studies.

**Methods:** The version 1 release contains WGS data derived from 3,941 participants from 4 cohorts. Samples underwent joint genotyping by the TOPMed Freeze 9 Variant Calling Pipeline. We performed descriptive analyses of these WGS data using the AMP PD platform.

**Results:** The clinical diagnosis of participants in version 1 release includes 2,005 idiopathic PD patients, 963 healthy controls, 64 prodromal subjects, 62 clinically diagnosed PD subjects without evidence of dopamine deficit (SWEDD) and 705 participants of genetically enriched cohorts carrying PD risk associated *GBA* variants or *LRRK2* variants in which 304 were affected. We did not observe a significant enrichment of pathogenic variants in the idiopathic PD group, but the polygenic risk score (PRS) was higher in PD both in non-genetically enriched cohorts and genetically enriched cohorts. The population analysis showed a correlation between genetically enriched cohorts and Ashkenazi Jewish ancestry.

**Conclusions:** We describe the genetic component of the AMP PD platform, a solution to democratise data access and analysis for the PD research community.

**(d) Financial Disclosure/CoI:** 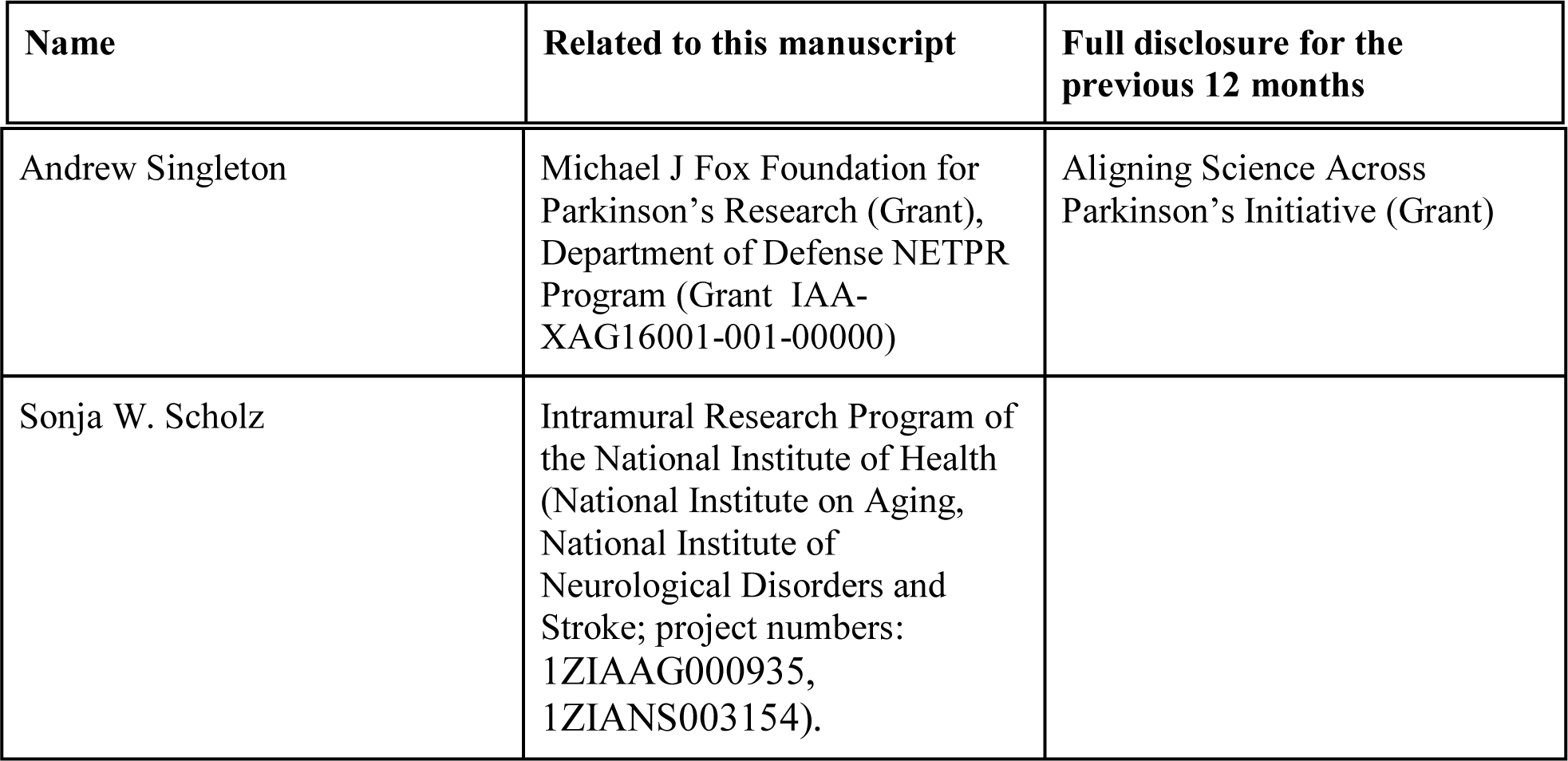

## Introduction

The genetic investigation of Parkinson’s disease (PD) has been a driving force in PD research over the last twenty years. Genetics serves to identify a starting point for the molecular and cellular processes that underlie disease. More recently, genetics has become part of an array of data types being used in an attempt to define disease at the individual level, with the aim of predicting who will get disease,^1^ when they will get it,^2^ and what their progression will look like.^3^ Ultimately genetics is a foundational part of the science that promises to reveal rational and viable targets for therapeutic intervention and to highlight the patients most suitable for each interventional strategy.^4,5^

This work has been both enabled and accelerated by the rapid development and adoption of methods for the generation and analysis of massive-scale genetic data. The application of genome-wide genotyping and whole-genome sequencing (WGS) has significantly altered the speed and potential of genetics research in PD, resulting in the rapid identification of genetic variability linked to disease. A critical challenge to the effective use of these data centers on data scale, production, and sharing. Genetics is expensive, can be challenging to analyze in a uniform way, and is often difficult to effectively share, both because of practical and regulatory reasons. Addressing these challenges promises to reduce duplicated effort, accelerate discovery, and democratize research.

A part of the Accelerating Medicines Partnership Parkinson’s Disease (AMP PD) project is centered on the development and deployment of a knowledge platform. It will present varied data relevant to PD. A large component of this data comes in the form of whole-genome sequence (WGS) that has been or will be generated across the Parkinson’s Progression Markers Initiative (PPMI), the Parkinson’s Disease Biomarkers Project (PDBP), the Harvard Biomarkers Study (HBS), BioFIND, the Study of Urate Elevation in Parkinson’s disease trial (SURE-PD), and the Safety, Tolerability and Efficacy Assessment of Dynacirc CR in Parkinson Disease (STEADY-PD) trial. At the time of writing, the AMP PD platform contains complete WGS data on 3,941 individuals from these studies.

The knowledge platform provides users with access to the genetic data and a space in which to perform analyses *in situ*, without download. The flexible nature of the underlying Google Cloud Platform architecture affords users the ability to quickly deploy compute resources to analyze genome-scale data. This platform also enables the user to deploy workflows that incorporate other data modalities, including phenotypic and transcriptomic datasets.

Here, we describe the data generation, processing, and quality control of these genetic data. We also provide the user with access to the workflows and pipelines that were used to perform these analyses, which can be copied or modified by the user. We provide a summary of the genetic characterization of these samples and provide corresponding annotated code to execute such analyses.

## Methods

### Cohorts

The release 1 (AMP PD v1_release) included 4 multicenter observational studies: BioFIND (https://biofind.loni.usc.edu), Harvard Biomarkers Study^5,6^ (HBS, https://www.bwhparkinsoncenter.org/biobank), Parkinson’s Disease Biomarker Program (PDBP, https://pdbp.ninds.nih.gov), and Parkinson’s Progression Markers Initiative (PPMI, https://www.ppmi-info.org). Participants’ clinical information and genetic samples were obtained under appropriate written consent and with local institutional and ethical approvals. The details of these studies can be obtained from the AMP PD website (https://amp-pd.org) and each study website. The data from SURE-PD and STEADY-PD are being processed for the next release.

### Data Flow Overview

The sample quality control steps and the released data are outlined in Figure 1. AMP PD requires quality control checks for all release-bound data at the sequencing facility first, to ensure minimum quality controls are met for individual samples before being transferred to AMP PD. For the flagship AMP PD data release version 1, all WGS samples (n=4,067) were processed from fastq and vetted through a series of independent genomic quality control (QC) checks and interdependent multi-modal QC checks. Samples passing all QC checks were processed using the Broad joint discovery pipeline and annotated with Variant Effect Predictor (VEP) (n=3,074), ^6^ or TOPMed variant calling pipeline (n=3,941) (https://www.nhlbiwgs.org). For each type of QC test, a plan was created through a collaboration of the AMP PD WGS working group and contributors from the NIH/NIA/LNG, wherein each described discrete checks and threshold values required of passing samples. These QC tests are outlined below and the scripts are shared in the AMP PD workspace with AMP PD useers (https://app.terra.bio/#workspaces/fc-amp-pd-alpha/AMP%20PD%20WGS%20QC%20Collaboration). During QC test execution, failing samples were noted against each discrete quality control check, so that each test result could be evaluated independently. This approach enabled the Transcriptomics working group and Clinical Data Harmonization working group to consider the impact of each WGS QC check to their constituent QC processes. The overall QC results for the AMP PD release could be interpreted without ambiguity as to which QC check resulted in the exclusion of a participant sample, whether the exclusion arose from a QC test in the WGS, Transcriptomic, or Clinical Data Harmonization working group.

**Figure 1.**
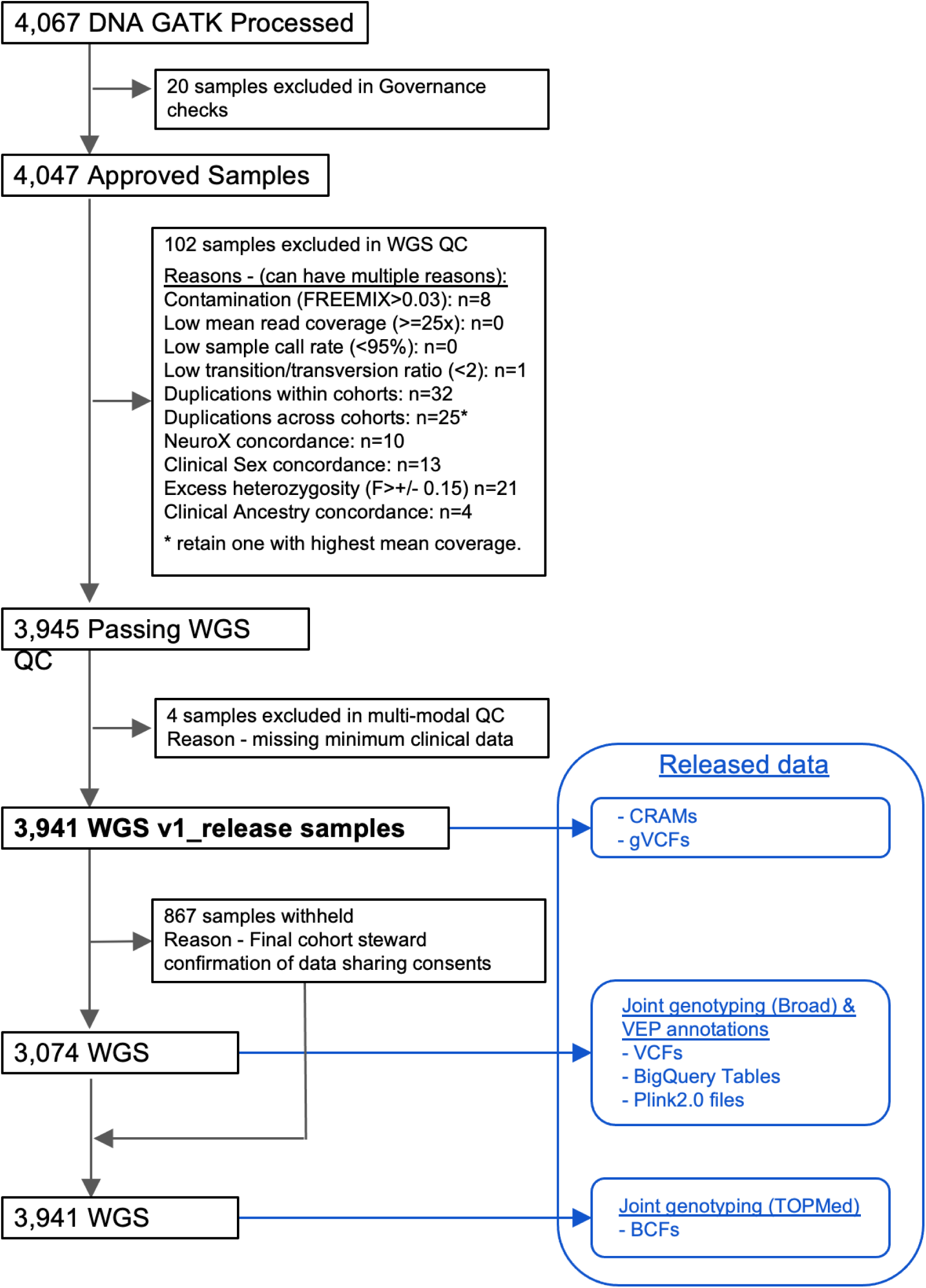
WGS Sample Flowchart WGS, whole genome sequencing; QC, quality control.

### DNA sequencing and variants calling

DNA samples were derived from the participants’ whole blood specimens and sequenced using Illumina HiSeq X Ten platform at Macrogen Corp or the Uniformed Services University of Health Sciences. Paired-end 300-400bp reads were processed in accordance with the functional equivalence pipeline^7^ implemented in the Broad Institute. Alignment and variant calling were against the GRCh38DH reference genome.

### WGS Quality Controls

AMP PD v1_release Quality Control tests include Governance checks that required contributing cohort stewards to identify participants that could be released by the AMP PD program in a formal artifact, the Subject Master List. Each cohort steward affirmed the identifiers to be used as the basis for AMP PD participant identifiers were free of Personal Identifiable Information (PII) and could not be deconstructed or reconstructed to reveal PII. Cohort stewards from contributing studies consented to modification of these identifiers to allow AMP PD to adapt them to a naming convention that was constructed and agreed to by participating members of the AMP PD working groups. We used two alphabets as a cohort identifier while a following 4 digit number to distinguish the participants. (e.g., BF-0011). This uniform naming convention was then adopted by each AMP PD working group and threaded through all data types to achieve a uniform representation of the participant in all filenames and file contents, across WGS, Transcriptomics, and Clinical Data records. The Governance QC check enabled the release of consistently named data and confirmed consented cohort participants.

The WGS working group prepared a plan for testing WGS samples for contamination, quality, duplicates across studies, duplicates within studies, and concordance with clinical and with pre-existing NeuroX genotyping array platform data.^8^ These tests were broken into discrete QC checks that were defined in great detail, documented, and executed by contributing experts from the NIH/NIA/LNG. The complete analysis resulted in a recommendation to the WGS Working Group for each QC check as to whether AMP PD should exclude a sample from the release, include the sample but withhold from joint genotyping results, or flag the sample as problematic for downstream consideration by the investigating end-user. QC checks for duplicates resulted in exclusion of all but one sample from joint genotyping, whereby the sample of higher mean coverage was selected for inclusion and all other samples were identified as duplicates in a release artifact and queryable table. Samples that passed WGS QC were further evaluated against other AMP PD data types. While WGS QC test results primarily informed downstream Transcriptomics tests, Clinical data QC tests were bi-directional. As the Clinical Data Harmonization group defined criteria for minimum clinical data, participant records were thus excluded during clinical QC, resulting in the exclusion of WGS samples (n=4). The Clinical data QC test for sample data asserts that no WGS or Transcriptomics data can be released without matching clinical participant data.

### Joint genotyping

The first set of Joint Genotyped variants consisted of 3,074 samples and was published by AMP PD in November, 2019. Joint Genotyping was run on Terra and used the Broad Institute’s joint discovery pipeline (workflow and fixed inputs can be found on GitHub, https://github.com/amp-pd/amp-pd-workflows). The Joint Genotyped VCF files were then run through the VEP using the annotations feature of the <u>Variant Transforms tool</u> from Google Cloud (https://github.com/googlegenomics/gcp-variant-transforms). The VEP database used is version 91 of homoserines, GRCh38. The 3,074 Joint Genotyped and annotated variants are made available in four different forms: per-chromosome gzipped VCFs, Plink 1.9 files, Plink 2.0 files, and as a table in Google BigQuery. The VCFs were loaded to BigQuery using the <u>vcf_to_bq</u> command of Variant Transforms.

More recently, we published all 3,941 samples in the release version 1 jointly genotyped by TOPMed Freeze 9 Variant Calling Pipeline (The web-page under preparation. The previous versions were described at https://www.nhlbiwgs.org/data-sets). The AMP PD samples were combined with 143,415 samples sequenced in the NHLBI TOPMed program, 60,540 samples sequenced in the NHGRI Centers for Common Disease Genomics (CCDG) program, 15,042 sequenced samples from NIA-NINDS studies and 2,504 samples from the 1000 Genomes Project Phase 3, deeply sequenced by the New York Genome Center. The genotypes for only the AMP PD samples were returned to AMP PD. Variant functional annotation is provided from snpEff 4.3t (build 2017-11-24 10:18),^9^ using the GRCh38.86 database. Statistically phased haplotypes using Eagle 2.4 (Dec 13, 2017)^10^ will be provided when they are ready.

### Descriptive analysis

We provide a descriptive analysis of baseline characteristics and of sequencing metrics. We summarized the carrier status of ClinVar “pathogenic” variants^11^ for autosomal dominant PD genes. To determine “pathogenic” variants, we applied two criteria. One derived a variant only annotated as “pathogenic” while the other included a wider set of variants that had at least one annotation such as “likely_pathogenic” or “pathogenic” among multiple annotations (pathogenic+). For autosomal recessive genes, we additionally considered loss of function variants (LoF). The LoF variants were defined as having “HIGH” impact consequences determined by VEP annotation which includes transcript ablation, splice acceptor variant, splice donor variant, stop gained frameshift variant, stop lost, start lost, and transcript amplification.^6^

The population structure of the participants was analyzed using HapMap samples of European, Asian, and African continental ancestry.^12^ We merged the study data with these referencing data and conducted a principal components analysis. Each continental-level ancestry was determined by mean ± 6 standard deviations from the reference panel. We also referenced genotyping array data from GSE23636 at Gene Expression Omnibus to identify the Ashkenazi Jewish population in the study.^13^ For participants of European descent, we calculated the polygenic risk score (PRS) using the weights of 90 significant variants from the recent meta-analysis of PD GWAS^1^ and conducted a descriptive analysis of PRS scores per study arm.

### Data availability

All data processing was conducted on the Google Cloud Platform. Processing/analysis scripts were provided at the related workspaces for reference. (Accessible for AMP PD users) The resulting CRAM files, VCFs and jointly genotyped data (BCF, VCF, PLINK and BigQuery format) are available through the AMP PD.

## Results

Table 1 shows the baseline characteristics and the sample-level sequencing quality metrics. Among 3,941 participants, there were 2,005 participants with idiopathic PD and 963 controls from idiopathic case-control cohorts. 705 participants were from the genetically enriched cohorts (the genetic cohort or the genetic registry of PPMI) of which 304 were affected and the rest were unaffected. These PPMI genetically enriched cohorts are individuals who are specifically recruited for their genetic status and include carriers of *LRRK2* p.G2019S, *GBA* p.N370S, *SNCA* p.A53T. Other study arms included participants with prodromal symptoms (n=64), SWEDDs (n=62), and disease controls (patients with other neurological diseases, n=127). The sequencing metrics were compatible with recent genetic studies with the median/mean coverage between 33.3x and 35.0x.

**Table 1.** Whole genome sequenced participants

|  | Overall | BioFIND | HBS | PDBP | PPMI |
| --- | --- | --- | --- | --- | --- |
| Total N | 3941 | 172 | 867 | 1469 | 1433 |
| **<u>Gender and Age</u>** |  |  |  |  |  |
| Female, n (%) | 1725 (43.8) | 71 (41.3) | 372 (42.9) | 640 (43.6) | 642 (44.8) |
| Age at baseline, years old, mean (SD) | 63.5 (10.7) | 67.1 (6.9) | 66.1 (10.1) | 64.0 (10.0) | 61.1 (11.7) |
| **<u>Self-reported race</u>** |  |  |  |  |  |
| White, n (%) | 3726 (94.6) | 161 (93.6) | 844 (97.3) | 1397 (95.1) | 1324 (92.5) |
| Mixed ancestry, n (%) | 65 (1.6) | 2 (1.2) | 2 (0.2) | 6 (0.4) | 55 (3.8) |
| Black or African American, n (%) | 63 (1.6) | 3 (1.7) | 10 (1.2) | 32 (2.2) | 18 (1.3) |
| Asian, n (%) | 34 (0.9) | 1 (0.6) | 7 (0.8) | 16 (1.1) | 10 (0.7) |
| **<u>Study arms</u>** |  |  |  |  |  |
| Parkinson's disease, n (%) | 2005 (51.1) | 99 (57.6) | 640 (73.8) | 858 (58.9) | 408 (28.5) |
| Healthy control, n (%) | 963 (24.5) | 73 (42.4) | 227 (26.2) | 470 (32.3) | 193 (13.5) |
| Genetic Cohort PD, n (%) | 179 (4.6) |  |  |  | 179 (12.5) |
| Genetic Cohort Unaffected, n (%) | 222 (5.7) |  |  |  | 222 (15.5) |
| Genetic Registry PD, n (%) | 125 (3.2) |  |  |  | 125 (8.7) |
| Genetic Registry Unaffected, n (%) | 179 (4.6) |  |  |  | 179 (12.5) |
| Prodromal, n (%) | 64 (1.6) |  |  |  | 64 (4.5) |
| SWEDD, n (%) | 62 (1.6) |  |  |  | 62 (4.3) |
| Disease Control, n (%) | 127 (3.2) |  |  | 127 (8.7) |  |
| **<u>Variant metrics</u>** |  |  |  |  |  |
| MEAN_COVERAGE, median [Q1,Q3] | 33.9 [31.2,36.2] | 35.0 [34.1,35.7] | 33.4 [30.7,36.3] | 33.3 [30.8,36.3] | 34.2 [31.8,36.4] |
| MEDIAN_COVERAGE, median [Q1,Q3] | 34.0 [32.0,37.0] | 35.0 [35.0,36.0] | 34.0 [31.0,37.0] | 34.0 [31.0,37.0] | 35.0 [32.0,37.0] |
| READS/K, median [Q1,Q3] | 3519 [3254,3760] | 3629 [3552,3690] | 3489 [3198,3765] | 3446 [3194,3742] | 3565 [3322,3789] |
| AVG_DP, median [Q1,Q3] | 35.2 [32.6,37.6] | 36.3 [35.5,36.9] | 34.9 [32.0,37.7] | 34.5 [31.9,37.4] | 35.7 [33.2,37.9] |
| FREEMIX, median [Q1,Q3] | **0.0 [0.0,0.0]** | 0.0 [0.0,0.0] | 0.0 [0.0,0.0] | 0.0 [0.0,0.0] | 0.0 [0.0,0.0] |

PD, Parkinson's disease; SWEDD, Scan without evidence of dopamine deficit; AVG\_DP, Average depth.

Carriers of pathogenic/Lof familial PD genes were summarized in Table 2. BigQuery enabled us to derive these variants of interest immediately. The carriers of these variants for *GBA* and *LRRK2* were relatively frequent because of the recruiting strategy for the targeted genetic recruitment. In the non-genetically enriched cohorts, the carrier frequencies between cases and controls were not statistically significant except for that of pathogenic+ variants of *GBA* (carriers/all were 30/1365 in cases and 5/736 in controls, P = 0.0069 in Fisher’s exact test). We observed a relatively high number of *PRKN* pathogenic/LoF carriers compared with those of the other genes of interest. The majority of them (n=125) were the carriers of a *PRKN* intron variant rs9364644 (G>A,C). Although the VEP annotated it as a high impact variant (splice donor variant), the variant was not significantly enriched among cases in non-genetically enriched cohorts, and clinical significance was unclear. Twenty-seven *SNCA* pathogenic variant (p.A53T) carriers were all from the genetically enriched cohorts.

**Table 2.**
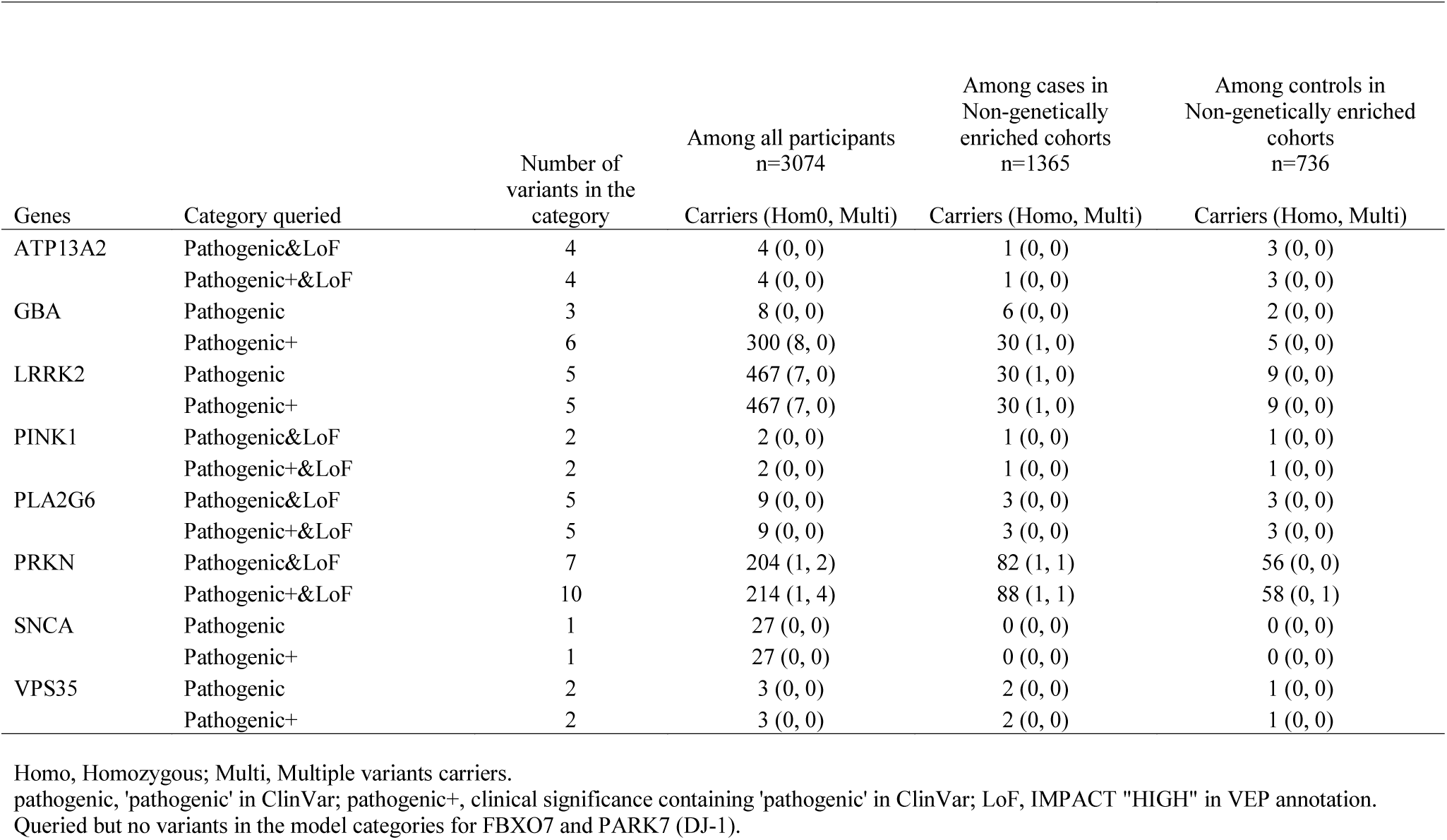
Pathogenic/LoF variants distribution of known PD genes

| Genes | Category queried | Number of<br>variants in the<br>category | Among all participants<br>n=3074 | Among cases in<br>Non-genetically<br>enriched cohorts<br>n=1365 | Among controls in<br>Non-genetically enriched<br>cohorts<br>n=736 |
| --- | --- | --- | --- | --- | --- |
|  |  |  | Carriers (Hom0, Multi) | Carriers (Homo, Multi) | Carriers (Homo, Multi) |
| ATP13A2 | Pathogenic&LoF | 4 | 4 (0, 0) | 1 (0, 0) | 3 (0, 0) |
|  | Pathogenic+&LoF | 4 | 4 (0, 0) | 1 (0, 0) | 3 (0, 0) |
| GBA | Pathogenic | 3 | 8 (0, 0) | 6 (0, 0) | 2 (0, 0) |
|  | Pathogenic+ | 6 | 300 (8, 0) | 30 (1, 0) | 5 (0, 0) |
| LRRK2 | Pathogenic | 5 | 467 (7, 0) | 30 (1, 0) | 9 (0, 0) |
|  | Pathogenic+ | 5 | 467 (7, 0) | 30 (1, 0) | 9 (0, 0) |
| PINK1 | Pathogenic&LoF | 2 | 2 (0, 0) | 1 (0, 0) | 1 (0, 0) |
|  | Pathogenic+&LoF | 2 | 2 (0, 0) | 1 (0, 0) | 1 (0, 0) |
| PLA2G6 | Pathogenic&LoF | 5 | 9 (0, 0) | 3 (0, 0) | 3 (0, 0) |
|  | Pathogenic+&LoF | 5 | 9 (0, 0) | 3 (0, 0) | 3 (0, 0) |
| PRKN | Pathogenic&LoF | 7 | 204 (1, 2) | 82 (1, 1) | 56 (0, 0) |
|  | Pathogenic+&LoF | 10 | 214 (1, 4) | 88 (1, 1) | 58 (0, 1) |
| SNCA | Pathogenic | 1 | 27 (0, 0) | 0 (0, 0) | 0 (0, 0) |
|  | Pathogenic+ | 1 | 27 (0, 0) | 0 (0, 0) | 0 (0, 0) |
| VPS35 | Pathogenic | 2 | 3 (0, 0) | 2 (0, 0) | 1 (0, 0) |
|  | Pathogenic+ | 2 | 3 (0, 0) | 2 (0, 0) | 1 (0, 0) |

Homo, Homozygous; Multi, Multiple variants carriers.
pathogenic, 'pathogenic' in ClinVar; pathogenic+, clinical significance containing 'pathogenic' in ClinVar; LoF, IMPACT "HIGH" in VEP annotation.
Queried but no variants in the model categories for FBXO7 and PARK7 (DJ-1).

The population analysis identified 95.3% (3,755/3,941) of the study participants were of European descent (Population plots in Supplemental Materials). Their PRS score distributions and other basic characteristics were summarized in Table 3. The mean PRS were significantly higher in PD cases (P = 3.5E-47, t-test) as well as SWEDDs (P = 0.033, t-test) than controls in the non-genetically enriched cohorts. The mean PRS of the affected were also significantly higher than the unaffected in the genetically enriched cohorts (P = 0.002, t-test). Participants in the genetically enriched cohorts had a higher PRS score than those in the non-genetically enriched cohorts (P < 1.0E-300, t-test). Indeed, the PRS scores showed distinguished distributions between the participants in the non-genetically enriched cohorts and the genetically enriched cohorts (Figure 2). This is due to the results of the recruiting strategy of these cohorts. Most of them carried the high-risk variant on *GBA, LRRK2*, or *SNCA* and when we re- calculated the PRS excluding 7 risk variants on these gene regions (rs114138760, rs35749011, rs76763715,rs34637584, rs76904798, rs5019538, and rs13117519), the polygenic risk scores (PRS83) distributions became similar (Figure 2). However, the mean PRS83 was still significantly different between the unaffected in the enriched cohorts and the healthy volunteers in the non-enriched cohorts. (P- value = 5.2E-5). When we calculated the effects of the risk variants on the PRS difference between the two arms, the variants with the largest 3 effect sizes were rs34637584 (*LRRK2* p.G2019S), rs76763715 (*GBA* p.N370S), and rs34311866 (*TMEM175* p.M393T). After adjusting for the three variants, PRS differences between the two arms were not significant anymore (P-value = 0.40, t-test). These variants are known to be enriched in the Ashkenazi Jewish population (AJ).^14^ We plotted the AJ reference with the study datasets, and it was indeed overlapped on a cluster of participants, especially those of genetically enriched cohorts. (Supplemental Materials). When we applied the cut-off of minimum PC3 among the AJ reference population to infer the AJ ancestry (PC3 = 0.156), the majority of the participants in the genetically enriched cohorts were inferred as AJ (Supplemental Materials).

**Table 3.** Cohort characteristics and polygenic risk score for European ancestry individuals

|  | Non-genetically enriched cohorts |  |  |  | Genetically enriched cohorts |  |
| --- | --- | --- | --- | --- | --- | --- |
|  | HC | PD | Prodromal | SWEDD | Unaffected | Affected |
| N | 905 | 1905 | 58 | 57 | 369 | 295 |
| Female, n (%) | 471 (52.0) | 675 (35.4) | 13 (22.4) | 21 (36.8) | 221 (59.9) | 153 (51.9) |
| Inferred AJ, n (%) | 73 (8.1) | 144 (7.6) | 2 (3.4) | 4 (7.0) | 263 (71.3) | 203 (68.8) |
| Age at baseline, years | 63.6 (10.8) | 64.7 (9.5) | 69.3 (5.9) | 60.9 (10.4) | 56.1 (12.7) | 65.5 (10.9) |
| **<u>Education levels</u>** |  |  |  |  |  |  |
| less than 12 years, n (%) | 13 (1.4) | 58 (3.0) | 13 (22.4) | 10 (17.5) | 17 (4.6) | 35 (11.9) |
| 12-16 years, n (%) | 659 (72.8) | 1405 (73.8) | 19 (32.8) | 32 (56.1) | 131 (35.5) | 136 (46.1) |
| Greater than 16 years, n (%) | 233 (25.7) | 440 (23.1) | 25 (43.1) | 15 (26.3) | 219 (59.3) | 123 (41.7) |
| **<u>Latest case/control status</u>** |  |  |  |  |  |  |
| case, n (%) | 3 (0.3) | 1887 (99.1) | 10 (17.2) | 50 (87.7) | 3 (0.8) | 292 (99.0) |
| control, n (%) | 896 (99.0) |  | 2 (3.4) | 1 (1.8) | 352 (95.4) | 1 (0.3) |
| Other (including prodromal state), n (%) | 6 (0.7) | 18 (0.9) | 46 (79.3) | 6 (10.5) | 14 (3.8) | 2 (0.7) |
| **<u>Polygenic risk scores</u>** |  |  |  |  |  |  |
| 90 common risk SNPs from Nalls et al (2019) | 0.0 (1.0) | 0.6 (1.1)*** | 0.1 (0.9) | 0.3 (0.9)* | 2.7 (1.9) | 3.2 (2.0)** |
| 83 SNPs from Nalls et al (2019) - excluding 7 variants in GBA, LRRK2 and SNCA regions | 0.0 (1.0) | 0.6 (1.0)*** | 0.1 (0.9) | 0.3 (0.9)* | 0.2 (1.0) | 0.5 (1.0)** |

Mean (SD) if not specified. Polygenic risk scores were standardized by the mean and the standard deviation of the scores in healthy controls in general cohorts. HC, Healthy controls or unaffected participants in genetic cohort/registry; PD, Parkinson's disease; SWEDD, Scan without evidence of dopaminergic deficit; AJ, Ashkenazi Jewish.
P-values for the score differences from the healthy controls (non-genetically enriched cohorts) or the unaffected (genetically enriched cohorts) were shown.
\*\*\*: $P < 0.001$ , \*\*: $P < 0.01$ , \*: $P < 0.05$ . (t-test).

**Figure 2.**
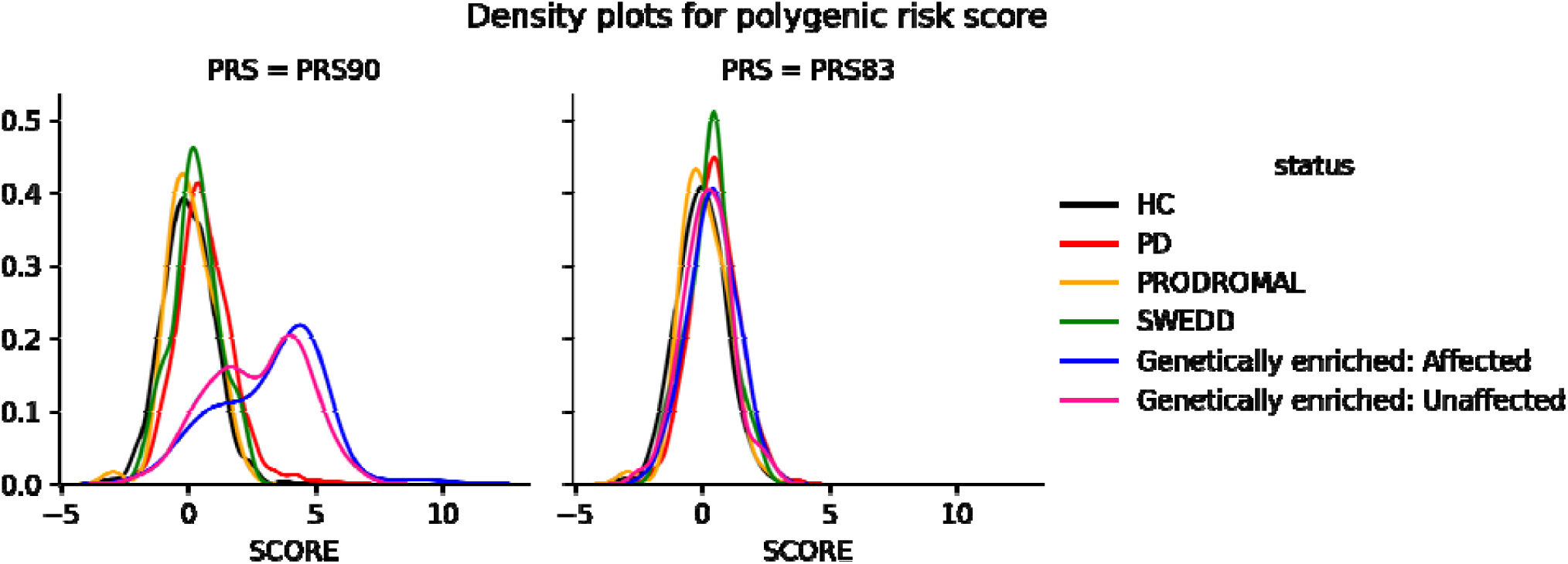
Density plots for polygenic risk score HC, healthy volunteers; PD, participants with Parkinson’s disease; SWEDD, Scan without evidence of dopamine deficit. PRS90 is a weighted sum of the independent risk loci reported in Nalls et al (2019). PRS83 is the same but removing the 7 variants in *GBA, LRRK2* and *SNCA* regions

## Discussion

Here we provide an overview of the DNA sequencing data that forms part of the first data release of the public-private partnership project AMP PD. The release version 1 contains WGS data from 3,941 participants. These data have undergone extensive quality control and standardized alignment and variant calling, including a single joint calling step. The data quality is high, enabling robust variant detection and calling across the full spectrum of variant frequencies.

We provide various formats of data: CRAMs, BCFs, VCFs, plink binary files, and BigQuery tables. As we demonstrated in the creation of Table 2, BigQuery allows rapid interrogation of the underlying data and retrieval of variants of interest. Tutorials for researchers not familiar with BigQuery are available on the AMP PD platform (https://app.terra.bio/#workspaces/fc-amp-pd-alpha/AMP%20PD%20-%20Workshop%20-%2020190508).

Our characterization of the WGS data available on the AMP PD platform as part of release 1 centered on topics that would likely be of broad interest to the users of these genetic data, or on issues that genetics could inform. The resource predominantly contains subjects of European Ancestry, and we believe that the genetically derived ancestry should be taken into account in many of the research questions that will be addressed with the AMP PD dataset, even those outside of genetics. Because of the design of the various contributing studies, a large number of subjects show Ashkenazi Jewish ancestry, driven by the preselection of genetic cases and the high number of *LRRK2* p.G2019S and *GBA* p.N370S carriers. The genetic characterization extends beyond the classification of these mutations to include a range of disease-linked mutations present in both cases and in as yet asymptomatic individuals. Again, we believe such information is likely to be key in potential clinical and biomarker analyses.

In addition to a characterization of disease-linked mutations, we also assessed the common genetic risk burden in these subjects. This calculation was based on the latest work identifying genetic risk loci in PD.^1^ The cases, as well as SWEDDs, carry a higher cumulative genetic burden of common PD risk variants compared to controls in the non-genetically enriched cohorts, as expected. Affected individuals also carry a higher burden of cumulative risk than unaffected individuals in the genetically enriched cohorts, in concordance with previous work.^15,16^ Importantly, the score distributions were substantially different across study arms, reflecting the different recruitment strategies of the study arms.

This project has a unique architecture. The AMP PD project provides an integrated analytical platform and much of the typical quality control and data processing that would be performed in WGS data has already been done to industry standards. Thus, while the underlying data are large, the most often used results and data forms have already been derived and can be readily accessed. Thus, researchers can concentrate on their own analyses without time-consuming logistics such as setting up and maintaining data sets and computational infrastructure. Transparency and extensibility are additional significant advantages of this project. Analyses using AMP PD data and the AMP PD platform are easily shared or copied and are inherently reproducible. The data processing scripts and analysis scripts used in this paper are shared in the AMP PD project and a cornerstone of the AMP PD philosophy is that other researchers are encouraged to share their processes, code, and results in the AMP PD analytical platform. We believe open science is the driving force of new discovery and the architecture of the project supports this approach.

A key aspect of the current AMP PD data is the harmonization of both a broad and deep range of data. A particular strength therefore of AMP PD will be the integrated analysis of these multi-modal data and most such analyses will include genetics. The available data types include transcriptomic data, biologic data from blood and cerebrospinal fluid, imaging summaries and detailed clinical phenotypes and test results. In addition, many of the data are available longitudinally. Immediate opportunities arise in the analysis of these data alone and integrated together. In the context of genetics one can imagine myriad uses, from adjustment for population structure, grouped analyses of clinical and biologic measures across suitably powered mutation types (both in cases and in asymptomatic carriers), and, importantly, analysis based on the varied burden of PD genetic risk score.

The primary limitation of the project from a purely genetic perspective is its size. Analyses on rare variants generally require a much larger sample size than that of common variants. A simulation reported that 5,000 cases and 5,000 controls are required to achieve the power of 0.8 for a burden test under the prior condition of the risk:non-risk variants ratio of 1:20 with a somewhat large relative risk of 5.^17^ Notably, current plans aim to substantially extend the number of genetically characterized subjects within AMP PD, thus the potential of this platform to support pure genetic discovery will improve with time.

Another limitation relates to the use of short-read sequencing technology. This method is less accurate and less powerful in detecting structural variants and tandem repeat variations.^18^ There are multiple tools proposed for calling structural variants and multi-algorithm consensus pipelines are proposed.^19^ However, it is difficult to capture breakpoints of structural variants containing repeats or embedded within repeats by aligning short-reads to a reference. Long-read sequencing technologies are expected to resolve these difficulties. Although it is still expensive and the error rate is high, it has been improving and it may be a promising future direction.

Notably, the accessible nature of AMP PD and its suitability for iterative and crowd-sourced analytical approaches means that as additional samples are added, and as novel analytical/processing strategies become available (for example calling structural variation) these can be rapidly deployed in AMP PD and this only needs to be done once to provide a standardized community resource.

In conclusion, we describe here the genetic arm of AMP PD, which includes a significant amount of raw and processed genetic data relevant to PD research and more broadly to neurodegenerative disease research. We believe this will be the foundation of a growing fund of genetic knowledge that will serve the PD research community.

## Acknowledgements

AMP PD – a public-private partnership – is managed by the FNIH and funded by Celgene, GSK, the Michael J. Fox Foundation for Parkinson’s Research, the National Institute of Neurological Disorders and Stroke, Pfizer, Sanofi, and Verily. This work was supported in part by the Intramural Research Program of the National Institute on Aging, National Institutes of Health, Department of Health and Human Services; project ZO1 AG000949. This research was also supported in part by the Intramural Research Program of the National Institute of Health (National Institute of Neurological Disorders and Stroke; project number: 1ZIANS003154).

Data used in the preparation of this article were obtained from the AMP PD Knowledge Platform. For up-to-date information on the study, **https://www.amp-pd.org**. Clinical data and biosamples used in preparation of this article were obtained from the Fox Investigation for New Discovery of Biomarkers (BioFIND), the Harvard Biomarker Study (HBS), the Parkinson’s Progression Markers Initiative (PPMI), and the Parkinson’s Disease Biomarkers Program (PDBP). BioFIND is sponsored by The Michael J. Fox Foundation for Parkinson’s Research (MJFF) with support from the National Institute for Neurological Disorders and Stroke (NINDS). The BioFIND Investigators have not participated in reviewing the data analysis or content of the manuscript. For up-to-date information on the study, visit michaeljfox.org/biofind. The Harvard Biomarkers Study (HBS) is a collaboration of HBS investigators [full list of HBS investigator found at https://www.bwhparkinsoncenter.org/biobank/] and funded through philanthropy and NIH and Non-NIH funding sources. The HBS Investigators have not participated in reviewing the data analysis or content of the manuscript. PPMI – a public-private partnership – is funded by the Michael J. Fox Foundation for Parkinson’s Research and funding partners, including [list the full names of all of the PPMI funding partners found at www.ppmi-info.org/fundingpartners. The PPMI Investigators have not participated in reviewing the data analysis or content of the manuscript. For up-to-date information on the study, visit www.ppmi-info.org. Parkinson’s Disease Biomarker Program (PDBP) consortium is supported by the National Institute of Neurological Disorders and Stroke (NINDS) at the National Institutes of Health. A full list of PDBP investigators can be found at https://pdbp.ninds.nih.gov/policy. The PDBP Investigators have not participated in reviewing the data analysis or content of the manuscript. Participation by individuals employed by Data Tecnica International LLC was supported in part by a consulting contract between the National Institutes of Health (NIA / NINDS) and the company. Individuals employed by Data Tecnica International LLC report now conflict of interest relating to the work carried out in this report.

