## Supplemental Materials for "Accelerating Medicines Partnership: Parkinson’s Disease. Genetic Resource"

### Supplemental Figure 1. Population plots with references populations

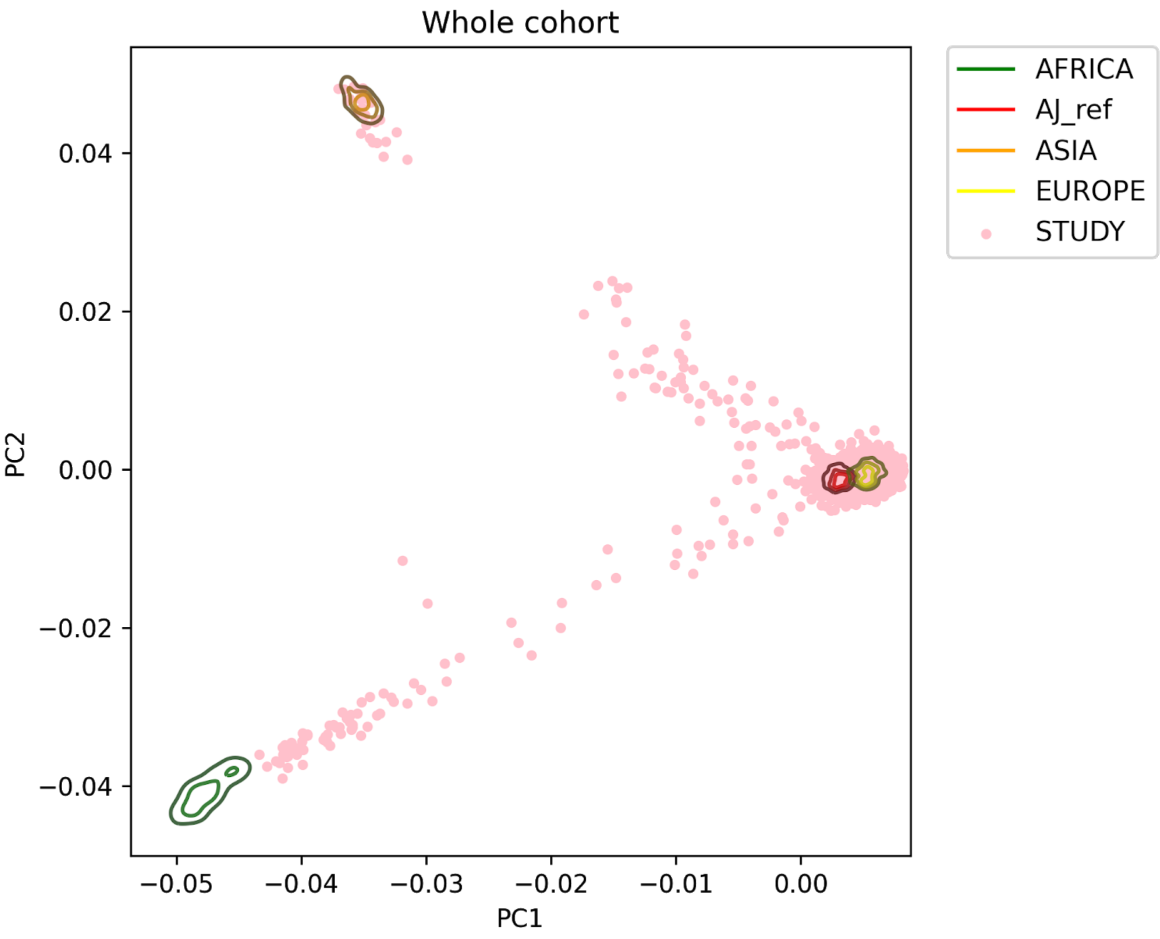

### Supplemental Figure 2. Genetic PC plots of European ancestry individuals

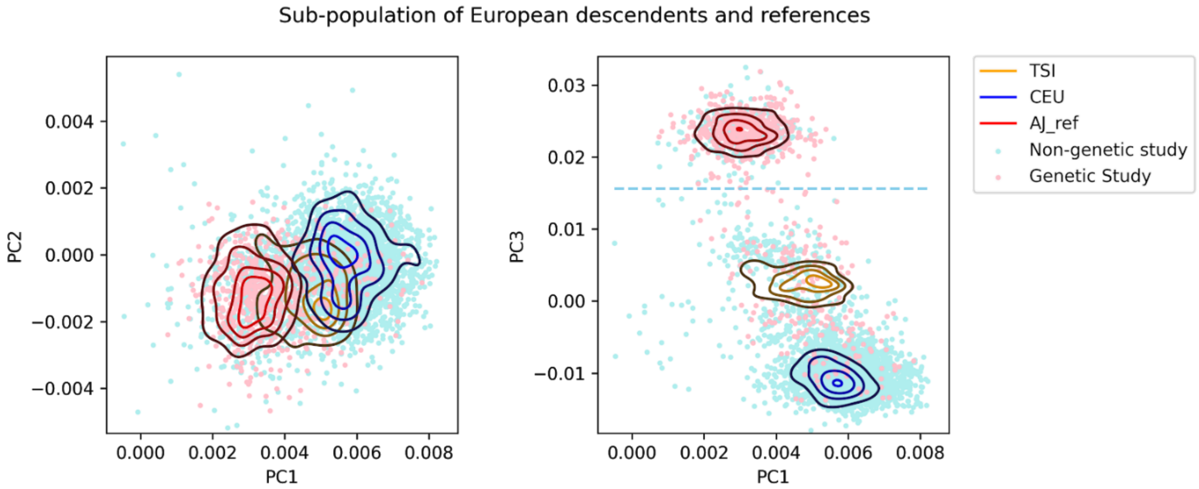


Blue dashed line represents the cut-off value to infer AJ population

AJ_ref, Ashkenazi Jewish reference panel; CEU Northern Europeans from Utah; TSI, Toscani from Italy.

Genetic study, genetically enriched cohorts; Non-genetic study, not genetically enriched cohorts.

### Supplemental Table. AJ percentage among Europeans

| Cohort | Category | N of Europeans | AJ % |
|---|---|---|---|
| BioFIND | | | |
| | HC | 70 | 14.3 |
| | PD | 99 | 16.2 |
| HBS | | | |
| | HC | 227 | 11.9 |
| | PD | 640 | 12.3 |
| PDBP | | | |
| | HC | 470 | 3.2 |
| | PD | 856 | 3.2 |
| PPMI Original Cohort | | | |
| | HC | 190 | 11.1 |
| | PD | 408 | 5.4 |
| | SWEDD | 61 | 6.6 |
| PPMI Prodromal Cohort | | | |
| | Prodromals | 61 | 3.3 |
| PPMI Genetic Cohort | | | |
| | HC_GC | 210 | 72.4 |
| | PD_GC | 178 | 53.9 |
| PPMI Genetic Registry | | | |
| | HC_GR | 164 | 67.7 |
| | PD_GR | 125 | 85.6 |
| All | | 3759 | 18.3 |

AJ, Ashkenazi Jewish; BF, BioFIND; PD, PDBP; PP, PPMI; HC, Healthy controls; PD, Parkinson's disease; GC, Genetic Cohort; GR, Genetic Registry.

### Group Authorships

| Name | Affiliation | AMP PD WGS Role |
|---|---|---|
| Andrew B Singleton | Center for Alzheimer's and Related Dementias, National Institute on Aging, Bethesda, MD, USA<br><br>Laboratory of Neurogenetics, National Institute on Aging, Bethesda, MD, USA | Co-chair |
| Ashutosh Pandey | GlaxoSmithKline, 9911 Belward Campus Dr, Rockville, MD 20850, USA | Member |
| Barry Landin | Technome, Herndon, VA, USA | Member |
| Bradford Casey | The Michael J. Fox Foundation for Parkinson's Research, New York, NY, USA | Member |
| Christine Swanson-Fischer | National Institute of Neurological Disorders and Stroke, National Institutes of Health, Bethesda, MD 20824, USA | Member |
| Clemens R. Scherzer | Harvard Medical School, Brigham and Women's Hospital, Boston, MA, USA | Member |
| David Pulford | GlaxoSmithKline, 9911 Belward Campus Dr, Rockville, MD 20850 | Member |
| David Vismer | Technome, Herndon, VA, USA | Member |
| Debra Babcock | National Institute of Neurological Disorders and Stroke, National Institutes of Health, Bethesda, MD 20824, USA | Member |
| Dena G Hernandez | Laboratory of Neurogenetics, National Institute on Aging, Bethesda, MD, USA | Member |
| Dinesh Kumar | Sanofi, Seattle, WA, USA | Member |
| Dongyu Liu | Sanofi, Seattle, WA, USA | Member |
| Eline Appelmans | 11400 Rockville Pike #600, North Bethesda, MD 20852, USA | Facilitator |
| Hampton L. Leonard | Data Tecnica International, Glen Echo, MD, USA<br><br>Center for Alzheimer's and Related Dementias, National Institute on Aging, Bethesda, MD, USA<br><br>Laboratory of Neurogenetics, National Institute on Aging, Bethesda, MD, USA | Member |
| Hirotaka Iwaki MD | Data Tecnica International, Glen Echo, MD, USA | Member |

| | | |
|---|---|---|
| | Center for Alzheimer's and Related Dementias, National Institute on Aging, Bethesda, MD, USA<br><br>Laboratory of Neurogenetics, National Institute on Aging, Bethesda, MD, USA | |
| J. Raphael Gibbs | Laboratory of Neurogenetics, National Institute on Aging, Bethesda, MD, USA | Member |
| Lynn Jakeman | National Institute of Neurological Disorders and Stroke, National Institutes of Health, Bethesda, MD 20824, USA | Member |
| Mahdiar Sadeghi | The American Genome Center, Uniformed Services University of the Health Sciences, Bethesda, MD, USA<br><br>Previously Sanofi, Seattle, WA, USA | Alumni |
| Mary B. Makarious | Laboratory of Neurogenetics, National Institute on Aging, Bethesda, MD, USA | Member |
| Margaret Sutherland | 801 Jefferson Avenue, Redwood, CA, USA<br><br>Previously National Institute of Neurological Disorders and Stroke, National Institutes of Health, Bethesda, MD 20824, USA | Alumni |
| Mark Frasier | The Michael J. Fox Foundation for Parkinson's Research, New York, NY, USA | Member |
| Matt Edwards | Verily Life Sciences, San Jose, CA, USA | Member |
| Matt Bookman | Verily Life Sciences, San Jose, CA, USA | Member |
| Meaghan Cogswell | Sanofi, Seattle, WA, USA | Member |
| Mike A. Nalls | Data Tecnica International, Glen Echo, MD, USA<br><br>Center for Alzheimer's and Related Dementias, National Institute on Aging, Bethesda, MD, USA<br><br>Laboratory of Neurogenetics, National Institute on Aging, Bethesda, MD, USA | Member |
| Robert Moccia | 1275 Pennsylvania Avenue NW STE 600, Washington, DC 20004, USA | Member |

| | | |
|---|---|---|
| Rosa Canet-Aviles | 35 Cambridge Park Dr Suite 200, Cambridge, MA 02140<br><br>Previously 11400 Rockville Pike #600, North Bethesda, MD 20852, USA | Alumni |
| Shameek Biswas | Bristol Myers Squibb, Seattle, WA, USA | Co-chair |
| Sonja W. Scholz | National Institute of Neurological Disorders and Stroke, Bethesda, MD, USA<br><br>Department of Neurology, Johns Hopkins University, Baltimore, MD, USA | Member |
| Srini Shankara | Sanofi, Seattle, WA, USA | Member |
| Xianjun Dong | Harvard Medical School, Brigham and Women's Hospital, Boston, MA, USA | Member |

### AMP PD consortium members

### Uniformed Services University of the Health Sciences Associates

| Name | Affiliation | Title and Role |
|---|---|---|
| Adelani Adeleye | Henry M. Jackson Foundation for the Advancement of Military Medicine, Inc., Bethesda, MD 20817 | System Administrator, implemented and conducted data transfer to NIH |
| Camille Alba | Henry M. Jackson Foundation for the Advancement of Military Medicine, Inc., Bethesda, MD 20817 | Research Assistant, managed project and sample tracking, supervised and conducted DNA quality control, library preparation, library quality control, sequencing for WGS |
| Dagmar Bacikova | Henry M. Jackson Foundation for the Advancement of Military Medicine, Inc., Bethesda, MD 20817 | Lab Manager, manages laboratory logistics and supplies for team personnel |
| Clifton L. Dalgard | Department of Anatomy, Physiology & Genetics, Uniformed Services University of the Health Sciences, Betheda, MD 20814 | Director, designed and directs all programmatic, laboratory and financial components for center activities |
| Daniel N. Hupalo | Henry M. Jackson Foundation for the Advancement of Military Medicine, Inc., Bethesda, MD 20817 | Computational Biologist, oversaw WGS data quality control and assembled data transfer selection and tracking |
| Elisa McGrath Martinez | Henry M. Jackson Foundation for the Advancement of Military Medicine, Inc., Bethesda, MD 20817 | Research Assistant, conducted library preparation, quality control, pooling and sequencing for WGS |
| Sraavya Polisetti | Henry M. Jackson Foundation for the Advancement of Military Medicine, Inc., Bethesda, MD 20817 | Research Technician, performed DNA quality assessment, quality control and sequencing for WGS |

| | | |
|---|---|---|
| John Rosenberger | Henry M. Jackson Foundation for the Advancement of Military Medicine, Inc., Bethesda, MD 20817 | Senior Research Assistant, oversaw sequencing platform performance for WGS |
| Anthony R. Soltis | Henry M. Jackson Foundation for the Advancement of Military Medicine, Inc., Bethesda, MD 20817 | Senior Computational Biologist, reviewed and evaluated WGS analysis pipeline and performance |
| Gauthaman Sukumar | Henry M. Jackson Foundation for the Advancement of Military Medicine, Inc., Bethesda, MD 20817 | Research Associate, conducted library preparation, quality control, pooling and sequencing for WGS |
| Miranda F. Tompkins | Henry M. Jackson Foundation for the Advancement of Military Medicine, Inc., Bethesda, MD 20817 | Research Technician, performed DNA quality assessment, quality control and sequencing for WGS |
| Meila Tuck | Henry M. Jackson Foundation for the Advancement of Military Medicine, Inc., Bethesda, MD 20817 | Research Technician, performed DNA quality assessment, quality control and sequencing for WGS |
| Coralie Violett | Henry M. Jackson Foundation for the Advancement of Military Medicine, Inc., Bethesda, MD 20817 | Scientist, conducted entry of subcohorts into project workload, initiated and tracked laboratory subprojects, reported subcohort quality control and completion |
| Matthew D. Wilkerson | Henry M. Jackson Foundation for the Advancement of Military Medicine, Inc., Bethesda, MD 20817 | Bioinformatics Director, designed and directs WGS analysis pipeline and performance |
| Xijun Zhang | Henry M. Jackson Foundation for the Advancement of Military Medicine, Inc., Bethesda, MD 20817 | Computational Biologist, designed and implements sequencing quality control dashboard and implemented panel-based cohort VCF generator |
